# Fine-tuned encoders, generative large language models and typed decisions for classifying findings in chest CT reports: paired English-Japanese internal testing and external testing on naturally written Japanese reports

**DOI:** 10.64898/2026.11.18.26368388

**Authors:** Yusuke Fukui, Sou Higashiyama, Yuhei Kawata, Yukihiro Nagatani, Harumi Iguchi, Yoshiyuki Watanabe

## Abstract

**Purpose:** To compare fine-tuned encoders, generative large language models (LLMs) and typed decisions (per-finding yes/no questions answered with a probability) for classifying 18 findings in chest CT reports, and to identify when typed decisions help.

**Methods:** The internal test set comprised 150 CT-RATE reports with paired Japanese translations at three quality levels; the external test set, 243 naturally written Japanese reports with human annotations (RR-Findings). We compared ModernBERT encoders fine-tuned on 20,444 reports, generative LLMs (gpt-6-luna, Qwen3.8-27B, MedGemma), and typed decisions with Jev, Luna Decisions, Gemini Decisions and Qwen. Thresholds fixed on 450 development reports were applied unchanged externally. The primary metric was micro-F1; case-level paired bootstrap 95% confidence intervals (CIs) were calculated for 15 internal comparisons.

**Results:** Encoders were best internally (English 0.9853; radiologist-checked Japanese 0.9576) and exceeded Jev (differences 0.0367, 95% CI 0.0258-0.0484; 0.0336, 0.0215-0.0464). Externally, Jev (0.8844) lay within the encoder seed range (0.8656-0.9144), exceeded the three generative conditions tested externally (best 0.8564), Luna Decisions (0.8177) and Gemini Decisions (0.7891), and had the smallest internal-to-external decrease (−0.0396). An encoder trained on 2,044 reports reached 0.8226 externally. Typed-versus-generated differences changed direction across models and test sets. At a fixed threshold of 0.5, Jev fell to 0.7753 externally.

**Conclusion:** Fine-tuned encoders performed best with ample in-distribution labels. Jev was an option with limited labels, but probability distributions differed across typed-decision services, so output probabilities and thresholds require validation on target reports.

## 1. Introduction

Structured information extracted from radiology reports supports case retrieval, quality control and labels for imaging models [1,2]. Rule-based labelers [1] and encoders fine-tuned on labeled reports [2] have been the standard approaches, and large language models (LLMs) now offer prompt-based extraction without task-specific training [3–5]. Typed decisions are now commercially available. Instead of generating text, the model answers typed questions about an input with probabilities, so a classifier can be defined at run time through question wording [6–8]. Typed decisions have been evaluated for general classification [9], medical question answering [10] and the factuality of radiology reports [11].

Comparisons of these approaches under identical conditions are scarce. In particular, we found no study that combined paired English–Japanese testing on the same cases and labels, external testing on reports from a different distribution, same-model comparisons of typed and generated outputs, and an analysis of calibration and threshold dependence.

We therefore compared fine-tuned encoders, generative LLMs and typed decisions for classifying 18 findings in chest CT reports with three aims: (1) to identify settings in which typed decisions outperform the other approaches; (2) to describe how each approach degrades under a small shift (translation quality of the same cases) and a large shift (naturally written Japanese reports); and (3) to test whether the performance of typed decisions comes from the format or from the implementation.

## 2. Materials and methods

### 2.1 Design and data

This retrospective comparison used public datasets only. Reporting followed TRIPOD-LLM [12] and CLAIM 2024 [13]. All settings were selected on development data independent of the test sets.

The internal test used CT-RATE [14] English reports and their Japanese translations in CT-RATE-JPN [15], which provides machine-translated (MT), resident-revised (revised) and radiologist-checked (checked) versions of the same reports (Table S13, Fig. S1). Both languages used the Findings and Impressions sections because CT-RATE’s automatic labeler was trained on annotations of their combined text [14]. Reference labels were the automatic labels distributed with CT-RATE, produced by a RadBERT classifier fine-tuned on 1,000 manually annotated reports [14], so the internal test measures agreement with an automatic labeler. The external test used RR-Findings [16] (revision 0911b306): 243 Japanese reports written by nine board-certified radiologists for 27 lung cancer cases, labeled for the same 18 findings by a radiology resident and reviewed by a board-certified radiologist, with final labels set by consensus [16]. The full report text was used as input.

### 2.2 Split and leakage control

Training data were restricted to 22,716 reports available in both languages. Reports whose Findings text also appeared in the test set were removed. Reports were split by connected components of patient identifiers and identical texts into 20,444 training and 2,272 development reports; 450 randomly selected development reports were used for model and threshold selection. The selection was not stratified by finding, but all 18 findings had positive labels (34–223 reports per finding). The internal test set comprised the 150 reports shared by both languages; their English and Japanese labels (2,700 each) were identical. Differences from the original Japanese ModernBERT study [16] are listed in Table S1.

### 2.3 Conditions

Table 1 lists the 16 English and 13 Japanese conditions. Encoders used ModernBERT [17], BioClinical ModernBERT [18] and a Japanese ModernBERT [19], each with an 18-output multi-label head. Generative conditions used gpt-6-luna [20] (Luna) with and without reasoning, Qwen3.8-27B [21] with and without thinking, and MedGemma [22,23] (English only), each zero-shot and with eight examples; each returned the 18 findings as 0/1 under a strict JSON schema. Typed decisions used Jev (jev-1.14.0) [6], the Decisions API of gpt-6-luna (Luna Decisions) [7] and Gemini Decisions (gemini-decision-1) [8], each sending the 18 questions in one request. A typed decision was also applied to Qwen, using the normalized probability of the first “Yes” or “No” token for each finding. Luna Decisions and the Qwen typed decision share their model with the corresponding generation; Jev, Luna Decisions and Gemini Decisions are three typed-decision services.

**Table 1.** Conditions.

| Approach | Condition | Model and setting | English | Japanese |
|---|---|---|---|---|
| Fine-tuned encoder | ModernBERT-base / -large | BioClinical-ModernBERT-base / -large | ✓ | — |
| Fine-tuned encoder | Japanese ModernBERT | modernbert-ja-130m | — | ✓ |
| Generation | Luna, zero-/few-shot, reasoning off/on | gpt-6-luna, reasoning effort none/medium | ✓ | ✓ |
| Generation | Qwen, zero-/few-shot, thinking off/on | Qwen3.8-27B | ✓ | ✓ |
| Generation | MedGemma, zero-/few-shot | medgemma-27b-text-it | ✓ | — |
| Typed decision | Jev | jev-1.14.0, 18 questions per request | ✓ | ✓ |
| Typed decision | Luna Decisions | gpt-6-luna, 18 predicates per request | ✓ | ✓ |
| Typed decision | Gemini Decisions | gemini-decision-1, 18 yes/no questions per request | ✓ | ✓ |
| Typed decision | Qwen typed decision | Qwen3.8-27B, one request per finding | ✓ | ✓ |

For Luna generation, reasoning effort was none with temperature 0 or medium with temperature omitted. Qwen generation used the sampling settings recommended in the model card [21]: with thinking off, temperature 0.7, top-p 0.8, top-k 20, min-p 0 and presence penalty 1.5; with thinking on (reasoning effort medium), temperature 1.0, top-p 0.95, top-k 20, min-p 0 and presence penalty 0. Both used repetition penalty 1. MedGemma used greedy decoding (temperature 0, top-p 1) [23]. The Qwen typed decision used unfiltered first-token log probabilities at temperature 1 (Methods S1).

Prompts and eight few-shot examples (seed 42) were fixed before testing and were not tuned on development or test performance (Methods S2). Inference ran between 19 October and 22 November 2026 (UTC; model versions in Table S14).

Encoders were trained with three seeds (42, 123 and 456) and three learning rates; epoch and learning rate were chosen by micro-F1 at a fixed threshold of 0.5 on the 450 development reports, before fitting per-finding thresholds. All other conditions except Luna Decisions and Gemini Decisions were run three times. Point estimates are means over seeds or runs. Two runs each of Luna Decisions and Gemini Decisions gave identical evaluation metrics, so one run of each was used (Methods S1).

### 2.4 Thresholds

For conditions returning probabilities, 18 per-finding thresholds maximizing joint micro-F1 on the 450 development reports were frozen before testing. English thresholds were fitted on English development reports; Japanese thresholds were fitted on machine-translated development reports, the same text type used to train the Japanese encoder. In the primary analysis, the Japanese thresholds were applied unchanged to all three Japanese translation versions and to the external naturally written Japanese reports. Results at a fixed threshold of 0.5 were computed from the same probabilities. Generative conditions used their 0/1 outputs directly.

### 2.5 Metrics and statistics

The primary metric was micro-F1 over all 18 findings. Secondary metrics were macro-F1, precision, recall, specificity, Matthews correlation coefficient, exact match, AUROC, mean average precision, Brier score, log loss and expected calibration error [24]. Only one decision was unavailable: Luna Decisions refused to answer for pleural effusion in one English report; it was counted as negative, with its probability missing.

For 15 internal comparisons (Table 4), case-level paired bootstrap 95% CIs used 10,000 resamples [25], averaging run- or seed-specific estimates within each resample. Bonferroni-adjusted (99.67%) intervals were a sensitivity analysis. A condition was called higher or lower than another when the 95% CI excluded zero. External results received no CIs because the 243 reports derive from 27 cases without a published row-to-case mapping. External comparisons were described by whether run or seed ranges overlapped.

### 2.6 Training-data size

In a post hoc analysis, each encoder was retrained on 1/2, 1/10 and 1/100 of the training reports (10,222, 2,044 and 204), sampled by multi-label stratification [26] and nested within each seed, with the same development set and selection rules.

## 3. Results

### 3.1 Internal test

Encoders were best in both languages (Table 2). ModernBERT-large exceeded Jev in English (difference 0.0367, 95% CI 0.0258–0.0484), and the Japanese encoder exceeded Jev on the radiologist-checked version (0.0336, 0.0215–0.0464). Among conditions without task-specific training, Jev exceeded the best generative condition in English (0.0157, 0.0014–0.0294; the Bonferroni interval included zero) but not in Japanese (−0.0018, −0.0175 to 0.0136). Luna Decisions scored lowest in English (0.8381); one finding in one report was refused. Gemini Decisions scored 0.9398 in English and 0.9114 in checked Japanese. Translation quality changed micro-F1 by at most 0.012 (Table S2).

**Table 2.** Internal test (150 reports; development thresholds)

| **Condition** | **Runs** | **English micro-F1** | **Range** | **English macro-F1** | **Checked micro-F1** | **Range** | **Checked macro-F1** |
|---|---|---|---|---|---|---|---|
| ModernBERT-large, fine-tuned | 3 | 0.9853 | 0.9848–0.9856 | 0.9848 | — | — | — |
| ModernBERT-base, fine-tuned | 3 | 0.9790 | 0.9753–0.9822 | 0.9786 | — | — | — |
| Qwen typed decision | 3 | 0.9508 | — | 0.9518 | 0.9284 | — | 0.9249 |
| Jev (typed decision) | 3 | 0.9486 | 0.9482–0.9491 | 0.9545 | 0.9239 | 0.9222–0.9264 | 0.9213 |
| Gemini Decisions (typed decision) | 1 | 0.9398 | — | 0.9425 | 0.9114 | — | 0.9156 |
| Qwen generation, few-shot, thinking on | 3 | 0.9329 | 0.9305–0.9371 | 0.9197 | 0.9179 | 0.9146–0.9231 | 0.9048 |
| Luna generation, few-shot, no reasoning | 3 | 0.9322 | 0.9309–0.9334 | 0.9269 | 0.9258 | 0.9243–0.9269 | 0.9170 |
| MedGemma generation, few-shot | 3 | 0.9301 | — | 0.9321 | — | — | — |
| Qwen generation, few-shot, thinking off | 3 | 0.9291 | 0.9248–0.9323 | 0.9183 | 0.9180 | 0.9170–0.9195 | 0.9088 |
| Luna generation, few-shot, reasoning | 3 | 0.9273 | 0.9256–0.9298 | 0.9123 | 0.9230 | 0.9214–0.9243 | 0.9054 |
| Qwen generation, zero-shot, thinking on | 3 | 0.9258 | 0.9242–0.9274 | 0.9139 | 0.9108 | 0.9086–0.9146 | 0.9014 |
| Luna generation, zero-shot, no reasoning | 3 | 0.9254 | 0.9229–0.9270 | 0.9260 | 0.9129 | 0.9118–0.9138 | 0.9186 |
| Qwen generation, zero-shot, thinking off | 3 | 0.9199 | 0.9191–0.9205 | 0.9131 | 0.9102 | 0.9062–0.9126 | 0.9068 |
| Luna generation, zero-shot, reasoning | 3 | 0.9183 | 0.9164–0.9197 | 0.9047 | 0.9067 | 0.9044–0.9088 | 0.8979 |
| MedGemma generation, zero-shot | 3 | 0.9075 | — | 0.9173 | — | — | — |
| Luna Decisions (typed decision) | 1 | 0.8381 | — | 0.8879 | 0.9120 | — | 0.9103 |
| Japanese ModernBERT, fine-tuned | 3 | — | — | — | 0.9576 | 0.9557–0.9599 | 0.9513 |

Micro-F1 and macro-F1 are means of values calculated separately for each seed or run; Luna Decisions and Gemini Decisions values are from one representative run each. Range: minimum–maximum over seeds or runs; — for single or identical runs. All English metrics: Table S3.

### 3.2 External test

All conditions declined externally (Table 3, Fig. 1). Jev (0.8844, range 0.8799–0.8903) lay within the encoder seed range (mean 0.8927, range 0.8656–0.9144) and exceeded the best generative condition (0.8564, range 0.8509–0.8624) and Luna Decisions (0.8177), with non-overlapping ranges. Jev had higher macro-F1 than the encoder (0.6410 vs 0.5711). Gemini Decisions scored 0.7891 externally. With lower label prevalence externally (0.130 vs 0.217 internally), conditions without task-specific training shifted toward positive calls (precision 0.6558–0.8163, recall 0.9554–0.9754). Jev had the smallest internal-to-external decrease (−0.0396; encoder −0.0649). With some seeds, the encoder predicted individual findings poorly, for example cardiomegaly and pulmonary fibrotic sequela (Table S5).

**Table 3.** External test (243 reports; thresholds frozen on CT-RATE)

| **Condition** | **Runs** | **Micro-F1** | **Range** | **Macro-F1** | **Precision** | **Recall** | **Specificity** | **MCC** | **AUROC** |
|---|---|---|---|---|---|---|---|---|---|
| Japanese ModernBERT, fine-tuned | 3 | 0.8927 | 0.8656–0.9144 | 0.5711 | 0.9246 | 0.8644 | 0.9891 | 0.8784 | 0.9713 |
| Jev (typed decision) | 3 | 0.8844 | 0.8799–0.8903 | 0.6410 | 0.8163 | 0.9648 | 0.9676 | 0.8695 | 0.9799 |
| Qwen generation, zero-shot, thinking on | 3 | 0.8564 | 0.8509–0.8624 | 0.6078 | 0.7761 | 0.9554 | 0.9588 | 0.8386 | — |
| Luna Decisions (typed decision) | 1 | 0.8177 | — | 0.6307 | 0.7039 | 0.9754 | 0.9388 | 0.8000 | 0.9146 |
| Qwen generation, zero-shot, thinking off | 3 | 0.8044 | 0.8038–0.8050 | 0.6246 | 0.6895 | 0.9654 | 0.9351 | 0.7848 | — |
| Gemini Decisions (typed decision) | 1 | 0.7891 | — | 0.6124 | 0.6643 | 0.9718 | 0.9267 | 0.7699 | 0.9559 |
| Qwen typed decision | 3 | 0.7844 | — | 0.6204 | 0.6568 | 0.9736 | 0.9241 | 0.7653 | 0.9723 |
| Luna generation, zero-shot, no reasoning | 3 | 0.7816 | 0.7773–0.7840 | 0.5830 | 0.6558 | 0.9671 | 0.9242 | 0.7615 | — |

Encoder AUROC: mean of three seeds. Macro-F1 includes three findings with no external positives (F1 of 0 by definition) and four with one positive each (Table S5).

**Table 4.** Main internal comparisons: micro-F1 difference (A − B) with 95% CIs.

| No. | Test set | A | B | A | B | Difference | 95% CI | 99.67% CI | 95% CI verdict |
|---|---|---|---|---|---|---|---|---|---|
| 1 | English | ModernBERT-large, fine-tuned | Jev (typed decision) | 0.9853 | 0.9486 | +0.0367 | +0.0258 to +0.0484 | +0.0210 to +0.0553 | A higher |
| 2 | Japanese checked | Japanese ModernBERT, fine-tuned | Jev (typed decision) | 0.9576 | 0.9239 | +0.0336 | +0.0215 to +0.0464 | +0.0161 to +0.0528 | A higher |
| 3 | English | Jev (typed decision) | Qwen generation, few-shot, thinking on | 0.9486 | 0.9329 | +0.0157 | +0.0014 to +0.0294 | −0.0061 to +0.0371 | A higher |
| 4 | Japanese checked | Jev (typed decision) | Luna generation, few-shot, no reasoning | 0.9239 | 0.9258 | −0.0018 | −0.0175 to +0.0136 | −0.0250 to +0.0218 | No difference |
| 5 | English | Jev (typed decision) | Luna Decisions (typed decision) | 0.9486 | 0.8381 | +0.1104 | +0.0936 to +0.1284 | +0.0848 to +0.1380 | A higher |
| 6 | Japanese checked | Jev (typed decision) | Luna Decisions (typed decision) | 0.9239 | 0.9120 | +0.0119 | −0.0002 to +0.0239 | −0.0061 to +0.0291 | No difference |
| 7 | English | Luna Decisions (typed decision) | Luna generation, zero-shot, no reasoning | 0.8381 | 0.9254 | −0.0873 | −0.1040 to −0.0722 | −0.1124 to −0.0655 | A lower |
| 8 | Japanese checked | Luna Decisions (typed decision) | Luna generation, zero-shot, no reasoning | 0.9120 | 0.9129 | −0.0008 | −0.0132 to +0.0109 | −0.0192 to +0.0173 | No difference |
| 9 | English | Qwen typed decision | Qwen generation, zero-shot, thinking off | 0.9508 | 0.9199 | +0.0308 | +0.0171 to +0.0445 | +0.0105 to +0.0522 | A higher |
| 10 | Japanese checked | Qwen typed decision | Qwen generation, zero-shot, thinking off | 0.9284 | 0.9102 | +0.0182 | +0.0060 to +0.0310 | +0.0002 to +0.0367 | A higher |
| 11 | Japanese checked | Japanese ModernBERT, fine-tuned (1/10 data) | Jev (typed decision) | 0.9299 | 0.9239 | +0.0059 | −0.0090 to +0.0215 | −0.0164 to +0.0293 | No difference |
| 12 | English | Jev (typed decision) | Gemini Decisions (typed decision) | 0.9486 | 0.9398 | +0.0088 | +0.0006 to +0.0174 | −0.0039 to +0.0212 | A higher |
| 13 | Japanese checked | Jev (typed decision) | Gemini Decisions (typed decision) | 0.9239 | 0.9114 | +0.0125 | +0.0001 to +0.0248 | −0.0058 to +0.0304 | A higher |
| 14 | English | Luna Decisions (typed decision) | Gemini Decisions (typed decision) | 0.8381 | 0.9398 | −0.1017 | −0.1185 to −0.0856 | −0.1283 to −0.0775 | A lower |
| 15 | Japanese checked | Luna Decisions (typed decision) | Gemini Decisions (typed decision) | 0.9120 | 0.9114 | +0.0006 | −0.0109 to +0.0124 | −0.0161 to +0.0176 | No difference |

Case-level paired bootstrap (10,000 resamples). The 99.67% CI is Bonferroni-adjusted for all 15 comparisons.

**Fig 1.**
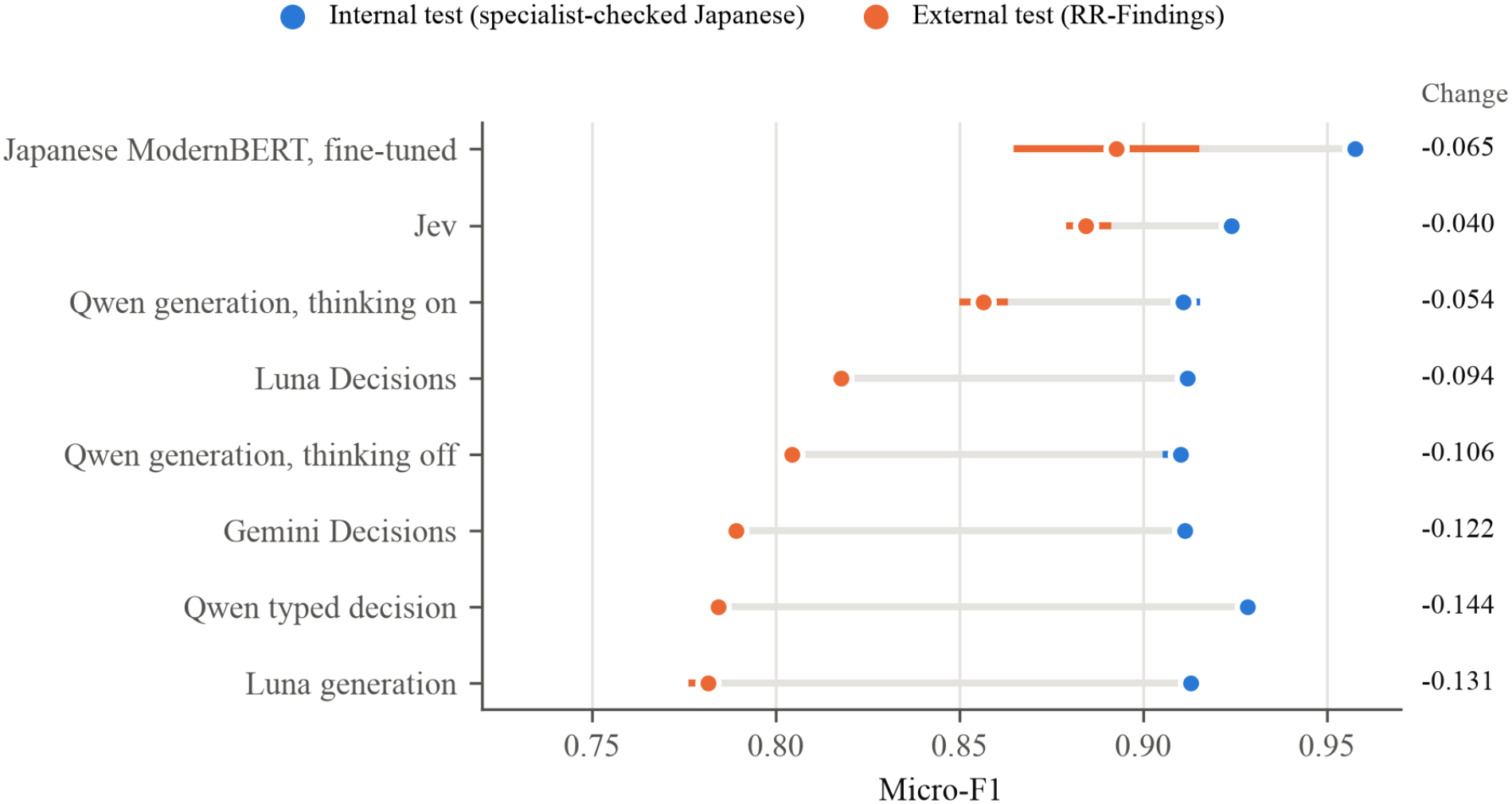
Micro-F1 on the internal test (radiologist-checked Japanese) and the external test for conditions evaluated on both. Bars show ranges over seeds or runs; the right column gives the change.

Encoder AUROC: mean of three seeds. Macro-F1 includes three findings with no external positives (F1 of 0 by definition) and four with one positive each (Table S5).

### 3.3 Typed-versus-generated and typed-decision service comparisons

The difference between typed and generated outputs reversed direction across models and test sets (Table 4, Table S4). On gpt-6-luna, Luna Decisions was lower than Luna generation in English (−0.0873, 95% CI −0.1040 to −0.0722), showed no difference in Japanese and was higher externally (+0.0361). On Qwen, the typed decision exceeded generation internally (English +0.0308; Japanese +0.0182) and was lower externally (−0.0200). Among the typed-decision services, Jev exceeded Luna Decisions in English (+0.1104, 0.0936–0.1284) and externally (+0.0667), with no difference in Japanese. Jev was higher than Gemini Decisions in English and checked Japanese by the 95% intervals, but both Bonferroni intervals included zero. Luna Decisions was lower than Gemini Decisions in English and showed no difference in checked Japanese. Discrimination and calibration metrics are reported in Table S7; probability distributions are shown in Fig. S2.

Case-level paired bootstrap (10,000 resamples). The 99.67% CI is Bonferroni-adjusted for all 15 comparisons.

### 3.4 Thresholds, calibration, efficiency and variability

The typed-decision services depended on per-finding thresholds: at 0.5, Jev fell by 0.0399 internally in English and by 0.1090 externally, whereas encoders changed by 0.0044 or less (Table S6). Externally, Luna Decisions and Gemini Decisions also decreased (−0.0400 and −0.0340). In the post hoc external threshold refit, Jev reached 0.9387 and the encoder 0.9292 on the same 243 reports used to fit the thresholds (Table S10). Expected calibration error was lower for encoders (external 0.0183) than for Jev (0.0968) (Table S7). Jev took about 0.17 s per report, compared with 2.0–5.7 s for Luna generation through the same type of API and 7.6–89.0 s for locally run generation (different environments; Table S8). Two runs each of Luna Decisions and Gemini Decisions gave identical evaluation metrics; MedGemma and the Qwen typed decision gave identical decisions across three runs. Jev and the remaining generative conditions changed 0.04–2.31% of decisions between runs, and the encoder seed range widened from 0.0042 internally to 0.0488 externally (Table S9).

### 3.5 Training-data size

With 2,044 training reports, the Japanese encoder showed no difference from Jev on the radiologist-checked version (0.9299 vs 0.9239; difference 0.0059, 95% CI −0.0090 to 0.0215) and scored 0.8226 externally, below Jev, with no seed above the upper end of Jev’s range (Fig. 2, Table S11). With 204 reports, it scored 0.6684 internally and 0.3747 externally. English encoders kept full-data performance down to 2,044 reports.

**Fig 2.**
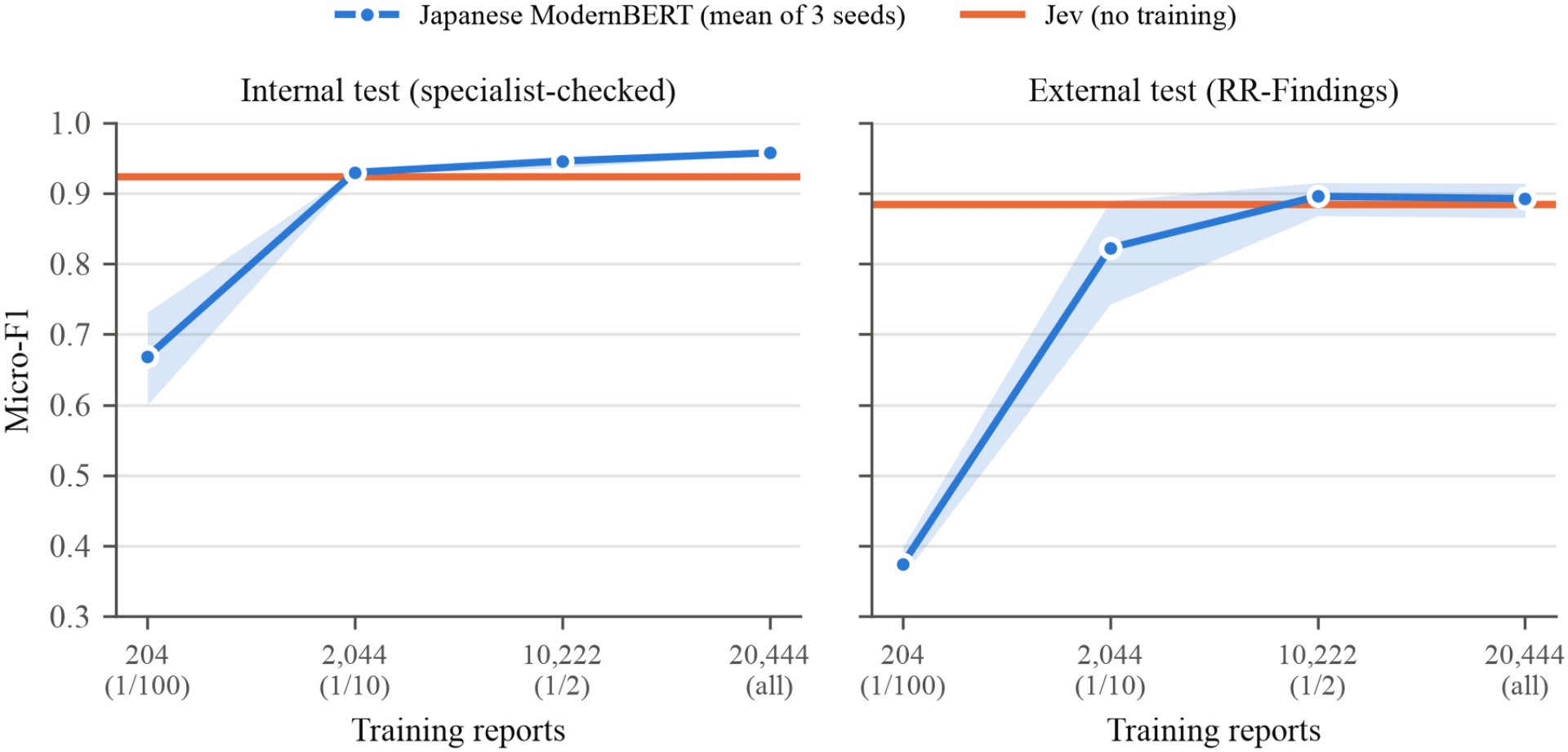
Japanese encoder micro-F1 by number of training reports (mean and range over three seeds) compared with Jev, which used no task-specific training (line: mean; band: range over three runs).

## 4. Discussion

### 4.1 Closeness to the training data

The choice among approaches depended less on output format than on how closely the training data matched the target reports and on whether labeled reports were available for setting thresholds.

Fine-tuned encoders were strongest when the test reports came from their training source. The internal reference labels were produced by an automatic labeler trained on the same report source [14], so an encoder trained on 20,444 of these labels could reproduce the labeler’s conventions, which no approach without task-specific training could do. This advantage did not depend on translation quality: the Japanese encoder remained the highest-scoring condition across all three translation versions (Table 2, Table S2). Within one report source, a fine-tuned encoder is therefore the most accurate choice.

The encoder performed less well when the report source changed. On naturally written Japanese reports, the encoder’s errors were mostly missed positives concentrated in a few findings and seeds, and its seed range widened (Table S5, Table S9). For cardiomegaly, the encoder ranked the positive reports above the negative ones but assigned them probabilities too low to cross its threshold (Table S5). Differences in writing style between machine-translated training text and naturally written Japanese reports are one possible explanation for the greater change in probabilities than in ranking. Jev, which was not trained on any of these reports, declined least and fell within the encoder seed range (Table 3, Fig. 1), and the encoder’s misses on sparse findings account for Jev’s higher macro-F1. The internal ranking of the approaches therefore did not carry over to a new report population.

### 4.2 Probability quality rather than format

Typed decisions did not consistently improve performance over generation. In same-model comparisons, the difference reversed between models and test sets: the Qwen typed decision exceeded Qwen generation internally but not externally, and Luna Decisions fell below Luna generation in English (Table 4, Table S4). Comparisons among Jev, Luna Decisions and Gemini Decisions also varied by language and report population.

Graded output alone did not explain these service differences. Both Jev and Gemini Decisions returned intermediate probabilities, whereas Luna Decisions concentrated its outputs at 0 and 1 (Fig. S2). Accordingly, a threshold of 0.5 lowered internal performance more for Jev and Gemini Decisions than for Luna Decisions, although externally Luna Decisions fell as much as Gemini Decisions (Table S6). Gemini Decisions had higher external AUROC and expected calibration error but lower log loss than Luna Decisions, so a single summary metric did not order all aspects of probability quality. Service selection therefore requires separate assessment of discrimination, calibration and classification at the intended thresholds.

### 4.3 Thresholds and training data

Jev’s preserved ranking (Table S7) made threshold placement a practical source of variation in external performance. With thresholds fitted on 450 development reports from another population, Jev exceeded the generative LLMs externally, whereas at a fixed threshold of 0.5 it fell below all three (Table 3, Table S6). Refitting the thresholds on the external reports raised its score further, although this post hoc in-sample value is optimistic (Table S10). Gemini Decisions also benefited from development thresholds rather than 0.5 (Table S6), and Deußer et al. [9] likewise reported gains from tuning Jev’s thresholds. When typed decisions such as Jev are used, fitting thresholds on a labeled sample from the target population is therefore a key step.

All conditions without task-specific training shifted toward positive calls on the external reports, whereas the encoder kept higher precision (Table 3). These conditions shared an instruction to count suspected or possible findings as present (Methods S2), whereas the encoder learned the treatment of such mentions from its training labels. This shared rule is a likely source of the shared shift, and locally fitted thresholds can partly correct it for Jev.

The encoder’s advantage also required ample training data. Trained on 2,044 reports, the Japanese encoder showed no internal difference from Jev but fell below it externally, with no seed above the upper end of Jev’s range, whereas Jev used 450 labeled reports only for thresholds (Fig. 2, Table S11). Less training data thus strengthened the encoder’s dependence on its training source, a weakness that internal testing did not reveal. When thousands of labeled reports resembling the target population cannot be prepared, Jev with thresholds fitted on a few hundred labeled reports is the more robust option across report populations (Fig. 2).

Generative LLMs need no thresholds and gave the highest external score among conditions that used no threshold-setting labels (Table 3, Table S6). Their shift toward positive calls and, with reasoning, their longer processing (Table S8) make them most suitable when no labeled reports are available.

### 4.4 Practical implications

The intended use is automated labeling of report text for case retrieval, quality control or research datasets, not decisions about individual patients, with performance verified by the user on labeled reports from the target population. The choice can follow two questions: whether the labeled training reports represent the target population, and whether labeled reports are available for threshold setting. With ample representative training data, a fine-tuned encoder gives the best agreement. With limited training data but a small labeled sample, Jev with locally fitted thresholds is preferable, and its performance at 0.5 should not be assumed to match that with fitted thresholds. Without any labels, a generative LLM is the remaining option. Deployment also differs. The encoders and Qwen run locally. The typed-decision services (Jev, Luna Decisions and Gemini Decisions) are readily usable for classification but send reports to commercial services, so data-protection requirements must be met. Furthermore, probability scales should not be assumed interchangeable: output probabilities and thresholds need validation on labeled reports from each target population. Because mean scores concealed poor predictions for individual findings by some encoder seeds, performance should be checked across seeds and findings.

### 4.5 Prior work

Earlier studies showed that generative LLMs can extract findings from free-text radiology reports without task-specific training [3–5], and typed decisions have been evaluated on medical question answering [10] and on the factuality of radiology reports [11]. For finding classification, this study adds that graded outputs alone do not explain service differences; discrimination, calibration and threshold-specific performance require separate evaluation across report populations. Jev’s short request time agrees with its reported speed advantage over LLM judges in a different task [27] (Table S8), and our encoder results agree with the Japanese ModernBERT study [16] (Table S12).

### 4.6 Limitations

First, internal labels were automatic, so internal results measure agreement with a labeler, not clinical accuracy. Second, the external set derives from 27 lung cancer cases without a row-to-case mapping, so external comparisons lack CIs and cannot establish equivalence between Jev and the encoder. The external input structure also differed from the internal set, and no external English set was available. Third, the internal models of Jev and Gemini Decisions are undisclosed. Fourth, the public test reports may have been in the models’ pretraining data, which was not tested (Table S14). Further limitations are listed in Limitations S1.

## 5. Conclusion

Fine-tuned encoders trained on ample in-distribution labels gave the highest agreement. Although Jev depended on thresholds and fell below the generative LLMs at 0.5, its strength appeared on reports from a different distribution. With thresholds fitted on only 450 labeled reports, it fell within the encoder seed range, declined least, exceeded the generative LLMs and an encoder trained on 2,044 reports, and ran faster than the generative LLMs. Luna Decisions and Gemini Decisions showed different patterns despite the shared typed format, and intermediate probabilities alone did not explain Jev’s advantage. Output probabilities and thresholds therefore require validation on labeled reports from each target population; whether fewer than 450 reports suffice remains to be tested.

## Declarations

### Ethics approval and consent

This study used only publicly available, de-identified datasets (CT-RATE, CT-RATE-JPN and RR-Findings) in accordance with their licenses, and no data were collected from human participants. Ethics committee approval and informed consent were therefore not required.

### Registration and protocol

The study was not registered. An analysis plan was prepared but not published; all settings are recorded in the code repository.

### Patient and public involvement

Patients and the public were not involved in the design, conduct, reporting or dissemination of this study.

### Data availability

CT-RATE (https://huggingface.co/datasets/ibrahimhamamci/CT-RATE, revision deeca4d8; CC BY-NC-SA 4.0), CT-RATE-JPN (https://huggingface.co/datasets/YYama0/CT-RATE-JPN, revision 41d80fdb; CC BY-NC-SA 4.0) and RR-Findings (https://huggingface.co/datasets/YYama0/RR-Findings, revision 0911b306; CC BY 4.0) are available from their distributors.

### Code availability

Code to prepare the data, run all conditions, set thresholds and reproduce the tables is available at https://github.com/shiga-med/ct-findings-typed-decision-benchmark under the MIT license; the version used in this study (v1.1.0) is archived at Zenodo (https://doi.org/10.5281/zenodo.24463329).

### Declaration of generative AI and AI-assisted technologies in the manuscript preparation process

During the preparation of this work the authors used Claude (Anthropic) to draft the English manuscript, generate tables from the study’s result files and check the reported numbers against those files. After using this tool, the authors reviewed and edited the content as needed and take full responsibility for the content of the published article.

## References

1. Irvin J, et al. CheXpert: a large chest radiograph dataset with uncertainty labels and expert comparison. Proc AAAI Conf Artif Intell. 2019;33(1):590–597. doi:10.1609/aaai.v33i01.3301590

2. Smit A, Jain S, Rajpurkar P, Pareek A, Ng AY, Lungren MP. Combining automatic labelers and expert annotations for accurate radiology report labeling using BERT. In: Proceedings of the 2020 Conference on Empirical Methods in Natural Language Processing (EMNLP). 2020:1500–1519. doi:10.18653/v1/2020.emnlp-main.117

3. Fink MA, et al. Potential of ChatGPT and GPT-4 for data mining of free-text CT reports on lung cancer. Radiology. 2023;308(3):e231362. doi:10.1148/radiol.231362

4. Le Guellec B, et al. Performance of an open-source large language model in extracting information from free-text radiology reports. Radiol Artif Intell. 2024;6(4):e230364. doi:10.1148/ryai.230364

5. Mukherjee P, Hou B, Lanfredi RB, Summers RM. Feasibility of using the privacy-preserving large language model Vicuna for labeling radiology reports. Radiology. 2023;309(1):e231147. doi:10.1148/radiol.231147

6. TypeSafe AI. Jev documentation: Models; Confidence; System One. https://docs.typesafe.ai/models, https://docs.typesafe.ai/confidence, https://docs.typesafe.ai/concepts/system-one (accessed 15 October 2026).

7. OpenAI. Decisions. API documentation. https://developers.openai.com/api/docs/guides/decisions (accessed 13 November 2026).

8. Choi M, Yu F. Gemini Decisions API: Fast, calibrated multimodal decisions. Google. 11 November 2026. https://blog.google/innovation-and-ai/technology/developers-tools/gemini-decisions-api/ (accessed 22 November 2026).

9. Deußer T, Ramamurthy R, Sparrenberg L, Sifa R. Evaluating and benchmarking the System One model Jev. arXiv:2609.37647, 2026 (preprint).

10. Madrid-García A, Merino-Barbancho B. Jev in medicine: a benchmark evaluation. arXiv:2609.34024, 2026 (preprint).

11. Huang J, Yang H, Ma X, et al. Can Jev judge radiology reports? Evaluating a System One model for clinical factuality. arXiv:2609.27607, 2026 (preprint).

12. Gallifant J, et al. The TRIPOD-LLM reporting guideline for studies using large language models. Nat Med. 2025;31(1):60–69. doi:10.1038/s41591-024-03425-5

13. Tejani AS, et al. Checklist for Artificial Intelligence in Medical Imaging (CLAIM): 2024 update. Radiol Artif Intell. 2024;6(4):e240300. doi:10.1148/ryai.240300

14. Hamamci IE, et al. Generalist foundation models from a multimodal dataset for 3D computed tomography. Nat Biomed Eng. 2026;10(8):1610–1628. doi:10.1038/s41551-025-01599-y

15. Yamagishi Y, et al. Development of a large-scale dataset of chest computed tomography reports in Japanese and a high-performance finding classification model: dataset development and validation study. JMIR Med Inform. 2025;13:e71137. doi:10.2196/71137

16. Yamagishi Y, et al. ModernBERT is more efficient than conventional BERT for chest CT findings classification in Japanese radiology reports. Sci Rep. 2026;16:15956. doi:10.1038/s41598-026-44292-z

17. Warner B, et al. Smarter, better, faster, longer: a modern bidirectional encoder for fast, memory efficient, and long context finetuning and inference. In: Proceedings of ACL 2025 (Volume 1: Long Papers). 2025:2526–2547. doi:10.18653/v1/2025.acl-long.127

18. Sounack T, Davis J, Durieux B, et al. BioClinical ModernBERT: a state-of-the-art long-context encoder for biomedical and clinical NLP. arXiv:2506.10896, 2025 (preprint).

19. SB Intuitions. modernbert-ja-130m. Hugging Face model card (sbintuitions/modernbert-ja-130m, revision 28c180b1).

20. OpenAI. GPT-6 Luna. Model documentation. https://developers.openai.com/api/docs/models/gpt-6-luna (accessed 21 October 2026).

21. Qwen Team. Qwen3.8-27B. Hugging Face model card (Qwen/Qwen3.8-27B, revision 1d4bf0f2).

22. Sellergren A, et al. An open vision-language model for diverse medical applications. Nat Med. 2026;32(10):3695–3704. doi:10.1038/s41591-026-04626-w

23. Google. medgemma-27b-text-it. Hugging Face model card (google/medgemma-27b-text-it, revision 5b667cf2).

24. Guo C, Pleiss G, Sun Y, Weinberger KQ. On calibration of modern neural networks. In: Proceedings of ICML 2017. PMLR. 2017;70:1321–1330.

25. Efron B, Tibshirani RJ. An Introduction to the Bootstrap. New York: Chapman & Hall; 1993.

26. Sechidis K, Tsoumakas G, Vlahavas I. On the stratification of multi-label data. In: Machine Learning and Knowledge Discovery in Databases (ECML PKDD 2011). Lecture Notes in Computer Science, vol 6913. Berlin: Springer; 2011:145–158. doi:10.1007/978-3-642-23808-6_10

27. Rao D, Callison-Burch C. Decision models vs. LLMs as rubric judges: cheaper, faster, and wrong in the same places. arXiv:2609.29769, 2026 (preprint).

