## Supplemental File for "Fine-tuned encoders, generative large language models and typed decisions for classifying findings in chest CT reports: paired English-Japanese internal testing and external testing on naturally written Japanese reports"

### Supplementary material

Fine-tuned encoders, generative large language models and typed decisions for classifying findings in chest CT reports. Performance values come from the saved predictions of the study and run-level values from the run logs; data characteristics come from the distributed metadata and published values from the cited studies.

#### Contents


- Methods S1. Details
  - Table S1. Data split and input
- Supplementary tables
  - Table S2. Internal test, Japanese: micro-F1 by translation version
  - Table S3. Internal test, English: all conditions
  - Table S4. Same-model typed-versus-generated comparisons and typed-decision service comparisons (micro-F1)
  - Table S5. Positives per finding, external F1 and encoder errors
  - Table S6. Development thresholds versus a fixed threshold of 0.5 (micro-F1)
  - Table S7. Discrimination and calibration
  - Table S8. Tokens and request time per report (first run of each condition unless noted)
  - Table S9. Variability over runs or seeds
  - Table S10. Post hoc threshold refitting and evaluation on the same external reports
  - Table S11. Encoders trained on reduced data
  - Table S12. Comparison with prior reports
- Limitations S1. Further limitations
- Methods S2. Data, reference standard, prompts and computing
  - Table S13. Characteristics of the data partitions
  - Fig. S1. Flow of reports
  - Table S14. Models, access and inference dates
- Probability distributions
  - Fig. S2. Returned positive-probability distributions


#### Methods S1. Details

**Split compared with the original Japanese ModernBERT study.**

**Table S1. Data split and input**

| Item | Original Japanese ModernBERT study [16] | This study: English | This study: Japanese |
|---|---|---|---|
| Training | 18,222 reports (80%) | 20,444 reports (about 90%) | The same 20,444 reports |
| Development | 4,556 reports (20%) | 2,272 reports (about 10%) | The same 2,272 reports |
| Selection set | — | 450 development reports | The same 450 reports, machine-translated |
| Test | 150 Japanese reports | 150 paired reports | The same 150 reports, three translation versions |
| Input | Findings only | Findings + Impressions | Findings + Impressions |

The 22,716 training-pool reports exclude 16 reports with missing text and 46 reports whose Findings text appears in the test set. Components of the split join reports that share a patient, an identical Findings text or an identical full text in either language. Each report has the same split and the same 18 labels in both languages; weights were trained separately for each language, and Japanese training used the machine-translated version. The 450-report subset was sampled from development reports with seed 42, without stratification by finding. All 18 findings had positive labels in this subset (34–223 reports per finding).

**Processing of the 18 findings.** Encoders: one forward pass with an 18-output multi-label head. Generation: one request returning the 18 findings as 0/1 under a strict JSON schema keyed by finding name (no probabilities). Typed decisions (Jev, Luna Decisions, Gemini Decisions and the Qwen typed decision): each finding was posed as the same yes/no question (Prompts by approach), and the returned probability of presence was converted to a decision with per-finding thresholds (Threshold rule). Jev, Luna Decisions and Gemini Decisions received the 18 questions on the same report in one request [6–8], identified by finding name (Jev, Luna Decisions) or by fixed identifiers finding_0–finding_17 (Gemini Decisions); the probability was the noul value for Jev and Gemini Decisions (not the separate confidence score) and the predicate probability for Luna Decisions. The Qwen typed decision used one request per finding (18 per report); the log probabilities of single-token spellings of "Yes" and "No" at the first output position were normalized to a probability, without token filtering. Reasoning settings were limited to each model's official switches (reasoning effort, thinking); MedGemma had none. One answer was generated per input.

**Encoder selection.** Each learning rate ($1\times10^{-5}$, $2\times10^{-5}$, $3\times10^{-5}$) and seed (42, 123, 456) was trained for up to five epochs. The epoch with the highest micro-F1 at 0.5 on the 450 development reports was selected (ties: earlier epoch), and the learning rate with the highest mean over seeds was chosen (ties: lower rate). Thresholds were then fitted on the same development probabilities.

**Threshold rule.** The 18 thresholds start at 0.5 and are updated by coordinate ascent on joint micro-F1, with candidates at every observed probability and at 0, 0.5 and 1; ties prefer the value closest to 0.5, then the higher value; a finding is positive when probability ≥ threshold. Thresholds were set separately for each condition and language. English thresholds were fitted on English development text; Japanese thresholds were fitted on machine-translated development text and shared across the three translation versions at test. In the primary analysis, Japanese thresholds were not refitted on the external reports.

**Repeated runs.** Jev, Luna and Qwen generation, MedGemma and the Qwen typed decision were run three times with the same inputs, model versions and thresholds. MedGemma generation used temperature 0. The Qwen typed decision used temperature 1 to obtain first-token log probabilities without token filtering; the probability was calculated from those log probabilities rather than the sampled token. The English Qwen generation runs used a per-request sampling seed. Two runs each of Luna Decisions and Gemini Decisions gave identical evaluation metrics, and one saved run of each was used. The Gemini Decisions repeat covered the evaluation reports only (600 internal and 243 external requests) with the thresholds of the first run.

**Wording of differences.** Internal comparisons with a 95% CI excluding zero are called higher or lower; otherwise "no difference" (not equivalence). Internal differences without CIs are reported descriptively. External differences are called higher or lower only when the ranges over runs or seeds do not overlap; otherwise "within the range". These external ranges do not account for case sampling variability.

#### Supplementary tables

##### Table S2. Internal test, Japanese: micro-F1 by translation version

| Condition | MT | Revised | Checked | Checked − MT |
|---|---|---|---|---|
| Japanese ModernBERT, fine-tuned | 0.9695 | 0.9611 | 0.9576 | −0.0119 |
| Qwen typed decision | 0.9323 | 0.9295 | 0.9284 | −0.0039 |
| Luna generation, few-shot, no reasoning | 0.9182 | 0.9247 | 0.9258 | +0.0075 |
| Jev (typed decision) | 0.9293 | 0.9269 | 0.9239 | −0.0054 |
| Luna generation, few-shot, reasoning | 0.9171 | 0.9255 | 0.9230 | +0.0060 |
| Qwen generation, few-shot, thinking off | 0.9153 | 0.9195 | 0.9180 | +0.0027 |
| Qwen generation, few-shot, thinking on | 0.9108 | 0.9196 | 0.9179 | +0.0072 |
| Luna generation, zero-shot, no reasoning | 0.9070 | 0.9120 | 0.9129 | +0.0058 |
| Luna Decisions (typed decision) | 0.9157 | 0.9184 | 0.9120 | −0.0037 |
| Gemini Decisions (typed decision) | 0.9154 | 0.9176 | 0.9114 | −0.0040 |
| Qwen generation, zero-shot, thinking on | 0.9117 | 0.9097 | 0.9108 | −0.0008 |
| Qwen generation, zero-shot, thinking off | 0.9098 | 0.9133 | 0.9102 | +0.0004 |
| Luna generation, zero-shot, reasoning | 0.9094 | 0.9086 | 0.9067 | −0.0027 |

##### Table S3. Internal test, English: all conditions

| Condition | Runs | Micro-F1 | Range | Macro-F1 | Exact match | Precision | Recall | Missing decisions |
|---|---|---|---|---|---|---|---|---|
| ModernBERT-large, fine-tuned | 3 | 0.9853 | 0.9848–0.9856 | 0.9848 | 0.8889 | 0.9809 | 0.9898 | 0 |
| ModernBERT-base, fine-tuned | 3 | 0.9790 | 0.9753–0.9822 | 0.9786 | 0.8578 | 0.9746 | 0.9835 | 0 |
| Qwen typed decision | 3 | 0.9508 | — | 0.9518 | 0.6933 | 0.9299 | 0.9727 | 0 |
| Jev (typed decision) | 3 | 0.9486 | 0.9482–0.9491 | 0.9545 | 0.6822 | 0.9277 | 0.9704 | 0 |
| Gemini Decisions (typed decision) | 1 | 0.9398 | — | 0.9425 | 0.6133 | 0.9213 | 0.9590 | 0 |
| Qwen generation, few-shot, thinking on | 3 | 0.9329 | 0.9305–0.9371 | 0.9197 | 0.6200 | 0.9410 | 0.9249 | 0 |
| Luna generation, few-shot, no reasoning | 3 | 0.9322 | 0.9309–0.9334 | 0.9269 | 0.6267 | 0.9303 | 0.9340 | 0 |
| MedGemma generation, few-shot | 3 | 0.9301 | — | 0.9321 | 0.5933 | 0.9075 | 0.9539 | 0 |
| Qwen generation, few-shot, thinking off | 3 | 0.9291 | 0.9248–0.9323 | 0.9183 | 0.5911 | 0.9231 | 0.9352 | 0 |
| Luna generation, few-shot, reasoning | 3 | 0.9273 | 0.9256–0.9298 | 0.9123 | 0.5933 | 0.9297 | 0.9249 | 0 |
| Qwen generation, zero-shot, thinking on | 3 | 0.9258 | 0.9242–0.9274 | 0.9139 | 0.5822 | 0.9150 | 0.9369 | 0 |
| Luna generation, zero-shot, no reasoning | 3 | 0.9254 | 0.9229–0.9270 | 0.9260 | 0.6067 | 0.9094 | 0.9420 | 0 |
| Qwen generation, zero-shot, thinking off | 3 | 0.9199 | 0.9191–0.9205 | 0.9131 | 0.5756 | 0.9026 | 0.9380 | 0 |
| Luna generation, zero-shot, reasoning | 3 | 0.9183 | 0.9164–0.9197 | 0.9047 | 0.5733 | 0.9005 | 0.9369 | 0 |
| MedGemma generation, zero-shot | 3 | 0.9075 | — | 0.9173 | 0.5133 | 0.9022 | 0.9130 | 0 |
| Luna Decisions (typed decision) | 1 | 0.8381 | — | 0.8879 | 0.1067 | 0.7527 | 0.9454 | 1 |

Missing decisions: findings without a usable answer, counted as negative (one Luna Decisions refusal for pleural effusion in an English report).

**Table S4. Same-model typed-versus-generated comparisons and typed-decision service comparisons (micro-F1)**

| Comparison (A vs B) | Test set | A | A range | B | B range | A − B |
|---|---|---|---|---|---|---|
| Same model (gpt-6-luna): typed vs generation | English | 0.8381 | — | 0.9254 | 0.9229–0.9270 | −0.0873 |
| Same model (gpt-6-luna): typed vs generation | Japanese MT | 0.9157 | — | 0.9070 | 0.9067–0.9073 | +0.0087 |
| Same model (gpt-6-luna): typed vs generation | Japanese revised | 0.9184 | — | 0.9120 | 0.9104–0.9135 | +0.0064 |
| Same model (gpt-6-luna): typed vs generation | Japanese checked | 0.9120 | — | 0.9129 | 0.9118–0.9138 | −0.0008 |
| Same model (gpt-6-luna): typed vs generation | External | 0.8177 | — | 0.7816 | 0.7773–0.7840 | +0.0361 |
| Same model (Qwen): typed vs generation | English | 0.9508 | — | 0.9199 | 0.9191–0.9205 | +0.0308 |
| Same model (Qwen): typed vs generation | Japanese MT | 0.9323 | — | 0.9098 | 0.9079–0.9117 | +0.0225 |
| Same model (Qwen): typed vs generation | Japanese revised | 0.9295 | — | 0.9133 | 0.9115–0.9147 | +0.0162 |
| Same model (Qwen): typed vs generation | Japanese checked | 0.9284 | — | 0.9102 | 0.9062–0.9126 | +0.0182 |
| Same model (Qwen): typed vs generation | External | 0.7844 | — | 0.8044 | 0.8038–0.8050 | −0.0200 |
| Typed-decision services: Jev vs Luna Decisions | English | 0.9486 | 0.9482–0.9491 | 0.8381 | — | +0.1104 |
| Typed-decision services: Jev vs Luna Decisions | Japanese MT | 0.9293 | 0.9259–0.9311 | 0.9157 | — | +0.0136 |
| Typed-decision services: Jev vs Luna Decisions | Japanese revised | 0.9269 | 0.9258–0.9282 | 0.9184 | — | +0.0085 |
| Typed-decision services: Jev vs Luna Decisions | Japanese checked | 0.9239 | 0.9222–0.9264 | 0.9120 | — | +0.0119 |
| Typed-decision services: Jev vs Luna Decisions | External | 0.8844 | 0.8799–0.8903 | 0.8177 | — | +0.0667 |
| Typed-decision services: Jev vs Gemini Decisions | English | 0.9486 | 0.9482–0.9491 | 0.9398 | — | +0.0088 |
| Typed-decision services: Jev vs Gemini Decisions | Japanese MT | 0.9293 | 0.9259–0.9311 | 0.9154 | — | +0.0139 |
| Typed-decision services: Jev vs Gemini Decisions | Japanese revised | 0.9269 | 0.9258–0.9282 | 0.9176 | — | +0.0093 |
| Typed-decision services: Jev vs Gemini Decisions | Japanese checked | 0.9239 | 0.9222–0.9264 | 0.9114 | — | +0.0125 |
| Typed-decision services: Jev vs Gemini Decisions | External | 0.8844 | 0.8799–0.8903 | 0.7891 | — | +0.0952 |
| Typed-decision services: Luna Decisions vs Gemini Decisions | English | 0.8381 | — | 0.9398 | — | −0.1017 |
| Typed-decision services: Luna Decisions vs Gemini Decisions | Japanese MT | 0.9157 | — | 0.9154 | — | +0.0003 |
| Typed-decision services: Luna Decisions vs Gemini Decisions | Japanese revised | 0.9184 | — | 0.9176 | — | +0.0008 |
| Typed-decision services: Luna Decisions vs Gemini Decisions | Japanese checked | 0.9120 | — | 0.9114 | — | +0.0006 |
| Typed-decision services: Luna Decisions vs Gemini Decisions | External | 0.8177 | — | 0.7891 | — | +0.0286 |

##### Table S5. Positives per finding, external F1 and encoder errors

| Finding | Internal positives (of 150) | External positives (of 243) | External F1: encoder | External F1: Jev | External F1: Qwen generation, thinking on | Encoder FN | Encoder FP | Encoder AUROC |
|---|---|---|---|---|---|---|---|---|
| Lymphadenopathy | 37 | 148 | 0.927 | 0.921 | 0.980 | 17.3 | 2.7 | 0.979 |
| Lung nodule | 82 | 121 | 0.849 | 0.949 | 0.894 | 19.3 | 18.7 | 0.912 |
| Pleural effusion | 19 | 67 | 0.987 | 0.992 | 0.995 | 1.7 | 0.0 | 1.000 |
| Lung opacity | 55 | 62 | 0.928 | 0.779 | 0.523 | 5.7 | 3.0 | 0.990 |
| Emphysema | 31 | 59 | 0.947 | 0.991 | 0.962 | 1.0 | 5.7 | 0.992 |
| Atelectasis | 49 | 51 | 0.959 | 0.990 | 0.987 | 4.0 | 0.0 | 0.986 |
| Pulmonary fibrotic sequela | 47 | 23 | 0.133 | 0.507 | 0.627 | 21.0 | 0.7 | 0.876 |
| Consolidation | 24 | 17 | 0.830 | 0.447 | 0.658 | 1.7 | 4.7 | 0.991 |
| Interlobular septal thickening | 7 | 8 | 1.000 | 0.961 | 0.980 | 0.0 | 0.0 | 1.000 |
| Cardiomegaly | 25 | 4 | 0.133 | 1.000 | 1.000 | 3.7 | 0.0 | 1.000 |
| Pericardial effusion | 12 | 4 | 0.952 | 1.000 | 1.000 | 0.3 | 0.0 | 1.000 |
| Arterial wall calcification | 49 | 1 | 0.244 | 0.667 | 0.000 | 0.3 | 3.0 | 0.988 |
| Bronchiectasis | 20 | 1 | 0.778 | 0.667 | 0.667 | 0.0 | 0.7 | 1.000 |
| Coronary artery wall calcification | 45 | 1 | 0.611 | 0.667 | 0.667 | 0.0 | 1.3 | 0.997 |
| Peribronchial thickening | 21 | 1 | 0.000 | 0.000 | 0.000 | 1.0 | 0.0 | 0.858 |
| Hiatal hernia | 24 | 0 | 0.000 | 0.000 | 0.000 | 0.0 | 1.0 | — |
| Medical material | 14 | 0 | 0.000 | 0.000 | 0.000 | 0.0 | 0.3 | — |
| Mosaic attenuation pattern | 25 | 0 | 0.000 | 0.000 | 0.000 | 0.0 | 0.0 | — |

Internal positives are for the 150 internal reports. F1 is 0 by definition for findings with no external positives. Encoder: mean of three seeds; others: mean of three runs. FN, false negatives; FP, false positives. For cardiomegaly, the four positive reports received encoder probabilities of 0.01–0.50 across seeds, all above every negative report (maximum 0.045) and all below 0.5.

##### Table S6. Development thresholds versus a fixed threshold of 0.5 (micro-F1)

| Condition | Test set | Development thresholds | Fixed 0.5 | Difference |
|---|---|---|---|---|
| ModernBERT-base, fine-tuned | English | 0.9790 | 0.9799 | +0.0008 |
| ModernBERT-large, fine-tuned | English | 0.9853 | 0.9850 | −0.0003 |
| Qwen typed decision | English | 0.9508 | 0.9148 | −0.0360 |
| Japanese ModernBERT, fine-tuned | Japanese checked | 0.9576 | 0.9586 | +0.0010 |
| Japanese ModernBERT, fine-tuned | Japanese MT | 0.9695 | 0.9691 | −0.0004 |
| Japanese ModernBERT, fine-tuned | Japanese revised | 0.9611 | 0.9615 | +0.0005 |
| Qwen typed decision | Japanese checked | 0.9284 | 0.8921 | −0.0362 |
| Qwen typed decision | Japanese MT | 0.9323 | 0.8962 | −0.0361 |
| Qwen typed decision | Japanese revised | 0.9295 | 0.8970 | −0.0326 |
| Luna Decisions (typed decision) | English | 0.8381 | 0.8173 | −0.0208 |
| Luna Decisions (typed decision) | Japanese checked | 0.9120 | 0.8970 | −0.0150 |
| Jev (typed decision) | English | 0.9486 | 0.9087 | −0.0399 |
| Jev (typed decision) | Japanese checked | 0.9239 | 0.8758 | −0.0481 |
| Jev (typed decision) | Japanese MT | 0.9293 | 0.8750 | −0.0544 |
| Jev (typed decision) | Japanese revised | 0.9269 | 0.8781 | −0.0488 |
| Luna Decisions (typed decision) | Japanese MT | 0.9157 | 0.8894 | −0.0263 |
| Luna Decisions (typed decision) | Japanese revised | 0.9184 | 0.8968 | −0.0215 |
| Luna Decisions (typed decision) | External | 0.8177 | 0.7777 | −0.0400 |
| Japanese ModernBERT, fine-tuned | External | 0.8927 | 0.8883 | −0.0044 |
| Jev (typed decision) | External | 0.8844 | 0.7753 | −0.1090 |
| Qwen typed decision | External | 0.7844 | 0.7829 | −0.0015 |
| Gemini Decisions (typed decision) | English | 0.9398 | 0.8975 | −0.0423 |
| Gemini Decisions (typed decision) | Japanese checked | 0.9114 | 0.8737 | −0.0377 |
| Gemini Decisions (typed decision) | Japanese MT | 0.9154 | 0.8711 | −0.0443 |
| Gemini Decisions (typed decision) | Japanese revised | 0.9176 | 0.8712 | −0.0464 |
| Gemini Decisions (typed decision) | External | 0.7891 | 0.7551 | −0.0340 |

##### Table S7. Discrimination and calibration

| Condition | Test set | AUROC | mAP | Brier | Log loss | ECE | Probability coverage | Probabilities of exactly 0 or 1, % |
|---|---|---|---|---|---|---|---|---|
| ModernBERT-base, fine-tuned | English | 0.9993 | 0.9973 | 0.0067 | 0.0260 | 0.0058 | 1.0000 | 0.0 |
| ModernBERT-large, fine-tuned | English | 0.9995 | 0.9980 | 0.0052 | 0.0224 | 0.0035 | 1.0000 | 0.1 |
| Qwen typed decision | English | 0.9934 | 0.9772 | 0.0300 | 0.1421 | 0.0237 | 1.0000 | 0.0 |
| Japanese ModernBERT, fine-tuned | Japanese checked | 0.9952 | 0.9831 | 0.0152 | 0.0673 | 0.0098 | 1.0000 | 0.0 |
| Japanese ModernBERT, fine-tuned | Japanese MT | 0.9985 | 0.9934 | 0.0103 | 0.0424 | 0.0056 | 1.0000 | 0.0 |
| Japanese ModernBERT, fine-tuned | Japanese revised | 0.9960 | 0.9843 | 0.0139 | 0.0612 | 0.0081 | 1.0000 | 0.0 |
| Qwen typed decision | Japanese checked | 0.9872 | 0.9671 | 0.0397 | 0.2092 | 0.0333 | 1.0000 | 0.0 |
| Qwen typed decision | Japanese MT | 0.9894 | 0.9628 | 0.0395 | 0.1962 | 0.0346 | 1.0000 | 0.0 |
| Qwen typed decision | Japanese revised | 0.9872 | 0.9672 | 0.0393 | 0.2061 | 0.0332 | 1.0000 | 0.0 |
| Luna Decisions (typed decision) | English | 0.9376 | 0.8631 | 0.0880 | 2.5847 | 0.0878 | 0.9996 | 94.7 |
| Luna Decisions (typed decision) | Japanese checked | 0.9592 | 0.8932 | 0.0435 | 1.0512 | 0.0407 | 1.0000 | 92.1 |
| Jev (typed decision) | English | 0.9931 | 0.9758 | 0.0342 | 0.1366 | 0.0529 | 1.0000 | 0.0 |
| Jev (typed decision) | Japanese checked | 0.9878 | 0.9564 | 0.0443 | 0.1786 | 0.0632 | 1.0000 | 0.0 |
| Jev (typed decision) | Japanese MT | 0.9894 | 0.9626 | 0.0435 | 0.1734 | 0.0635 | 1.0000 | 0.0 |
| Jev (typed decision) | Japanese revised | 0.9881 | 0.9585 | 0.0439 | 0.1772 | 0.0635 | 1.0000 | 0.0 |
| Luna Decisions (typed decision) | Japanese MT | 0.9601 | 0.8944 | 0.0447 | 0.9715 | 0.0421 | 1.0000 | 91.0 |
| Luna Decisions (typed decision) | Japanese revised | 0.9614 | 0.8993 | 0.0425 | 0.9903 | 0.0395 | 1.0000 | 92.1 |
| Luna Decisions (typed decision) | External | 0.9146 | 0.7292 | 0.0674 | 1.6171 | 0.0688 | 1.0000 | 93.0 |
| Japanese ModernBERT, fine-tuned | External | 0.9713 | 0.8156 | 0.0242 | 0.1078 | 0.0183 | 1.0000 | 0.0 |
| Jev (typed decision) | External | 0.9799 | 0.8131 | 0.0534 | 0.1935 | 0.0968 | 1.0000 | 0.0 |
| Qwen typed decision | External | 0.9723 | 0.8511 | 0.0588 | 0.2969 | 0.0585 | 1.0000 | 0.0 |
| Gemini Decisions (typed decision) | English | 0.9908 | 0.9661 | 0.0418 | 0.1761 | 0.0730 | 1.0000 | 0.0 |
| Gemini Decisions (typed decision) | Japanese checked | 0.9740 | 0.9380 | 0.0529 | 0.2158 | 0.0699 | 1.0000 | 0.0 |
| Gemini Decisions (typed decision) | Japanese MT | 0.9837 | 0.9477 | 0.0521 | 0.2087 | 0.0693 | 1.0000 | 0.0 |
| Gemini Decisions (typed decision) | Japanese revised | 0.9783 | 0.9501 | 0.0523 | 0.2115 | 0.0708 | 1.0000 | 0.0 |
| Gemini Decisions (typed decision) | External | 0.9559 | 0.7390 | 0.0766 | 0.3136 | 0.1325 | 1.0000 | 0.0 |

Probabilities of exactly 0 or 1: percentage of returned probabilities, pooled over runs or seeds; a decision at such a probability cannot be changed by moving the threshold.

Average precision was computed per finding as the sum over distinct probability values of the increase in recall multiplied by the precision at that value; cells with the same probability enter the ranking together as one threshold, so values do not depend on the order of tied cells (the definition used by scikit-learn's average_precision_score). Mean average precision is the mean over findings with at least one positive. AUROC assigned tied probabilities their mid-rank.

**Table S8. Tokens and request time per report (first run of each condition unless noted)**

| Condition | Test set | Requests | Input tokens/report | Output tokens/report | Seconds/report |
|---|---|---|---|---|---|
| Luna generation, zero-shot, no reasoning | English | 150 | 872 | 149 | 2.366 |
| Luna generation, few-shot, no reasoning | English | 150 | 5058 | 149 | 2.390 |
| Luna generation, zero-shot, reasoning | English | 150 | 872 | 369 | 4.256 |
| Luna generation, few-shot, reasoning | English | 150 | 5058 | 270 | 3.906 |
| Qwen generation, zero-shot, thinking off | English | 150 | 578 | 195 | 9.912 |
| Qwen generation, few-shot, thinking off | English | 150 | 4887 | 180 | 9.352 |
| Qwen generation, zero-shot, thinking on | English | 150 | 576 | 1196 | 59.522 |
| Qwen generation, few-shot, thinking on | English | 151 | 4917 | 1132 | 56.638 |
| MedGemma generation, zero-shot | English | 150 | 519 | 162 | 9.574 |
| MedGemma generation, few-shot | English | 150 | 4400 | 127 | 7.556 |
| Qwen typed decision | English | 2700 | 8141 | 18 | 3.418 |
| Luna generation, zero-shot, no reasoning | Japanese checked | 150 | 1240 | 149 | 2.087 |
| Luna generation, zero-shot, no reasoning | Japanese MT | 150 | 1252 | 149 | 2.046 |
| Luna generation, zero-shot, no reasoning | Japanese revised | 150 | 1240 | 149 | 2.090 |
| Luna generation, few-shot, no reasoning | Japanese checked | 150 | 6716 | 149 | 2.140 |
| Luna generation, few-shot, no reasoning | Japanese MT | 150 | 6728 | 149 | 2.003 |
| Luna generation, few-shot, no reasoning | Japanese revised | 150 | 6716 | 149 | 1.994 |
| Luna generation, zero-shot, reasoning | Japanese checked | 150 | 1240 | 381 | 4.379 |
| Luna generation, zero-shot, reasoning | Japanese MT | 150 | 1252 | 391 | 5.253 |
| Luna generation, zero-shot, reasoning | Japanese revised | 150 | 1240 | 373 | 4.719 |
| Luna generation, few-shot, reasoning | Japanese checked | 150 | 6716 | 276 | 5.655 |
| Luna generation, few-shot, reasoning | Japanese MT | 150 | 6728 | 287 | 4.904 |
| Luna generation, few-shot, reasoning | Japanese revised | 150 | 6716 | 282 | 3.088 |
| Qwen generation, zero-shot, thinking off | Japanese checked | 150 | 743 | 215 | 11.001 |
| Qwen generation, zero-shot, thinking off | Japanese MT | 150 | 750 | 216 | 11.044 |
| Qwen generation, zero-shot, thinking off | Japanese revised | 150 | 743 | 216 | 11.017 |
| Qwen generation, few-shot, thinking off | Japanese checked | 150 | 5056 | 170 | 8.882 |
| Qwen generation, few-shot, thinking off | Japanese MT | 150 | 5063 | 170 | 8.800 |
| Qwen generation, few-shot, thinking off | Japanese revised | 150 | 5056 | 170 | 8.735 |
| Qwen generation, zero-shot, thinking on | Japanese checked | 150 | 741 | 1761 | 88.041 |
| Qwen generation, zero-shot, thinking on | Japanese MT | 150 | 748 | 1782 | 88.967 |
| Qwen generation, zero-shot, thinking on | Japanese revised | 150 | 741 | 1742 | 86.998 |
| Qwen generation, few-shot, thinking on | Japanese checked | 150 | 5054 | 1374 | 68.825 |
| Qwen generation, few-shot, thinking on | Japanese MT | 150 | 5061 | 1361 | 68.171 |
| Qwen generation, few-shot, thinking on | Japanese revised | 151 | 5089 | 1440 | 72.056 |
| Qwen typed decision | Japanese checked | 2700 | 8853 | 18 | 3.623 |
| Qwen typed decision | Japanese MT | 2700 | 8966 | 18 | 3.632 |
| Qwen typed decision | Japanese revised | 2700 | 8851 | 18 | 3.621 |
| Jev (typed decision) | English | 150 | 2143 | 369 | 0.170 |

| Condition | Test set | Requests | Input tokens/report | Output tokens/report | Seconds/report |
|---|---|---|---|---|---|
| Jev (typed decision) | Japanese checked | 150 | 4319 | 369 | 0.174 |
| Jev (typed decision) | Japanese MT | 150 | 4338 | 369 | 0.171 |
| Jev (typed decision) | Japanese revised | 150 | 4319 | 369 | 0.169 |
| Luna Decisions (typed decision) | English | 150 | 4228 | 0 | 0.372 |
| Luna Decisions (typed decision) | Japanese checked | 151 | 5554 | 0 | 0.378 |
| Luna Decisions (typed decision) | Japanese MT | 150 | 5566 | 0 | 0.383 |
| Luna Decisions (typed decision) | Japanese revised | 150 | 5554 | 0 | 0.389 |
| Luna Decisions (typed decision) | External | 243 | 5222 | 0 | 0.385 |
| Japanese ModernBERT, fine-tuned (seed123) | External | 243 | — | — | 0.009 |
| Japanese ModernBERT, fine-tuned (seed42) | External | 243 | — | — | 0.011 |
| Japanese ModernBERT, fine-tuned (seed456) | External | 243 | — | — | 0.009 |
| Luna generation, zero-shot, no reasoning | External | 243 | 908 | 149 | 2.302 |
| Jev (typed decision) | External | 243 | 3933 | 369 | 0.169 |
| Qwen generation, zero-shot, thinking off | External | 243 | 516 | 214 | 10.895 |
| Qwen generation, zero-shot, thinking on | External | 243 | 514 | 1653 | 81.902 |
| Qwen typed decision | External | 4374 | 4753 | 18 | 2.281 |
| Gemini Decisions (typed decision) | English | 151 | — | — | 0.683 |
| Gemini Decisions (typed decision) | Japanese checked | 150 | 3058 | 0 | 0.652 |
| Gemini Decisions (typed decision) | Japanese MT | 150 | 3063 | 0 | 0.652 |
| Gemini Decisions (typed decision) | Japanese revised | 151 | — | — | 0.684 |
| Gemini Decisions (typed decision) | External | 243 | 2826 | 0 | 0.664 |

Request times of API and local GPU conditions were measured in different environments and are not directly comparable. For Jev on the revised Japanese version, the first run was slowed by a temporary interruption of communication and was excluded; its request time is the mean of the three adopted runs. For Gemini Decisions, token means are missing when a failed attempt lacked usage; request counts and times include those attempts. Recovered predictions do not remove the original failed attempts, whose usage and cost remain unknown. Successful requests were not resent.

##### Table S9. Variability over runs or seeds

| Condition | Test set | Source | n | Micro-F1 range | Changed decisions, % (min–max over pairs) |
|---|---|---|---|---|---|
| ModernBERT-base, fine-tuned | English | seeds | 3 | 0.0069 | 1.00–1.15 |
| ModernBERT-large, fine-tuned | English | seeds | 3 | 0.0008 | 0.63–0.74 |
| Luna generation, zero-shot, no reasoning | English | runs | 3 | 0.0040 | 0.41–0.48 |
| Luna generation, few-shot, no reasoning | English | runs | 3 | 0.0025 | 0.33–0.56 |
| Luna generation, zero-shot, reasoning | English | runs | 3 | 0.0033 | 0.41–0.59 |
| Luna generation, few-shot, reasoning | English | runs | 3 | 0.0042 | 0.30–0.63 |
| MedGemma generation, zero-shot | English | runs | 3 | 0.0000 | 0.00–0.00 |
| MedGemma generation, few-shot | English | runs | 3 | 0.0000 | 0.00–0.00 |
| Qwen typed decision | English | runs | 3 | 0.0000 | 0.00–0.00 |
| Japanese ModernBERT, fine-tuned | Japanese checked | seeds | 3 | 0.0042 | 1.26–1.96 |
| Japanese ModernBERT, fine-tuned | Japanese MT | seeds | 3 | 0.0044 | 1.07–1.22 |
| Japanese ModernBERT, fine-tuned | Japanese revised | seeds | 3 | 0.0024 | 1.22–1.74 |
| Luna generation, zero-shot, no reasoning | Japanese checked | runs | 3 | 0.0020 | 0.37–0.48 |
| Luna generation, zero-shot, no reasoning | Japanese MT | runs | 3 | 0.0006 | 0.52–0.59 |
| Luna generation, zero-shot, no reasoning | Japanese revised | runs | 3 | 0.0030 | 0.37–0.52 |

| Condition | Test set | Source | n | Micro-F1 range | Changed decisions, % (min–max over pairs) |
|---|---|---|---|---|---|
| Luna generation, few-shot, no reasoning | Japanese checked | runs | 3 | 0.0026 | 0.37–0.41 |
| Luna generation, few-shot, no reasoning | Japanese MT | runs | 3 | 0.0026 | 0.33–0.37 |
| Luna generation, few-shot, no reasoning | Japanese revised | runs | 3 | 0.0017 | 0.22–0.30 |
| Luna generation, zero-shot, reasoning | Japanese checked | runs | 3 | 0.0044 | 0.70–0.81 |
| Luna generation, zero-shot, reasoning | Japanese MT | runs | 3 | 0.0023 | 0.78–0.93 |
| Luna generation, zero-shot, reasoning | Japanese revised | runs | 3 | 0.0041 | 0.67–0.70 |
| Luna generation, few-shot, reasoning | Japanese checked | runs | 3 | 0.0029 | 0.41–0.67 |
| Luna generation, few-shot, reasoning | Japanese MT | runs | 3 | 0.0054 | 0.52–0.89 |
| Luna generation, few-shot, reasoning | Japanese revised | runs | 3 | 0.0083 | 0.56–0.78 |
| Qwen generation, zero-shot, thinking off | Japanese checked | runs | 3 | 0.0063 | 0.59–0.85 |
| Qwen generation, zero-shot, thinking off | Japanese MT | runs | 3 | 0.0038 | 0.67–0.89 |
| Qwen generation, zero-shot, thinking off | Japanese revised | runs | 3 | 0.0032 | 0.59–0.81 |
| Qwen generation, few-shot, thinking off | Japanese checked | runs | 3 | 0.0025 | 0.70–1.07 |
| Qwen generation, few-shot, thinking off | Japanese MT | runs | 3 | 0.0016 | 0.59–0.81 |
| Qwen generation, few-shot, thinking off | Japanese revised | runs | 3 | 0.0102 | 0.81–0.96 |
| Qwen generation, zero-shot, thinking on | Japanese checked | runs | 3 | 0.0060 | 0.89–1.11 |
| Qwen generation, zero-shot, thinking on | Japanese MT | runs | 3 | 0.0046 | 1.04–1.37 |
| Qwen generation, zero-shot, thinking on | Japanese revised | runs | 3 | 0.0032 | 0.78–1.07 |
| Qwen generation, few-shot, thinking on | Japanese checked | runs | 3 | 0.0085 | 1.19–1.33 |
| Qwen generation, few-shot, thinking on | Japanese MT | runs | 3 | 0.0030 | 1.33–1.56 |
| Qwen generation, few-shot, thinking on | Japanese revised | runs | 3 | 0.0051 | 1.07–1.41 |
| Qwen typed decision | Japanese checked | runs | 3 | 0.0000 | 0.00–0.00 |
| Qwen typed decision | Japanese MT | runs | 3 | 0.0000 | 0.00–0.00 |
| Qwen typed decision | Japanese revised | runs | 3 | 0.0000 | 0.00–0.00 |
| Qwen generation, zero-shot, thinking off | English | runs | 3 | 0.0014 | 0.52–0.74 |
| Qwen generation, few-shot, thinking off | English | runs | 3 | 0.0076 | 0.67–0.78 |
| Qwen generation, zero-shot, thinking on | English | runs | 3 | 0.0031 | 0.74–0.81 |
| Qwen generation, few-shot, thinking on | English | runs | 3 | 0.0067 | 1.00–1.11 |
| Jev (typed decision) | English | runs | 3 | 0.0009 | 0.04–0.15 |
| Jev (typed decision) | Japanese checked | runs | 3 | 0.0042 | 0.26–0.33 |
| Jev (typed decision) | Japanese MT | runs | 3 | 0.0052 | 0.30–0.52 |
| Jev (typed decision) | Japanese revised | runs | 3 | 0.0024 | 0.19–0.30 |
| Japanese ModernBERT, fine-tuned | External | seeds | 3 | 0.0488 | 1.78–3.18 |
| Luna generation, zero-shot, no reasoning | External | runs | 3 | 0.0067 | 0.53–0.57 |
| Jev (typed decision) | External | runs | 3 | 0.0104 | 0.48–0.85 |
| Qwen generation, zero-shot, thinking off | External | runs | 3 | 0.0012 | 1.62–1.87 |
| Qwen generation, zero-shot, thinking on | External | runs | 3 | 0.0115 | 2.08–2.31 |
| Qwen typed decision | External | runs | 3 | 0.0000 | 0.00–0.00 |

Luna Decisions and Gemini Decisions are not shown: two runs gave identical evaluation metrics, and one saved run of each was used (Methods S1).

##### Table S10. Post hoc threshold refitting and evaluation on the same external reports

| Condition | Runs | Internal (checked) | External, frozen thresholds | External, refit and evaluated on same data | Recovery |
|---|---|---|---|---|---|
| Luna Decisions (typed decision) | 1 | 0.9120 | 0.8177 | 0.8356 | +0.0179 |
| Japanese ModernBERT, fine-tuned | 3 | 0.9576 | 0.8927 | 0.9292 | +0.0365 |
| Jev (typed decision) | 3 | 0.9239 | 0.8844 | 0.9387 | +0.0544 |
| Qwen typed decision | 3 | 0.9284 | 0.7844 | 0.9097 | +0.1253 |

The 243 external reports were used both to fit the thresholds (from their labels and predicted probabilities) and to calculate micro-F1 with those thresholds; no separate test set was used. These in-sample values are optimistic and do not estimate performance after threshold setting on a separate sample.

##### Table S11. Encoders trained on reduced data

| Encoder | Test set | Training data | Micro-F1 | Range | Macro-F1 |
|---|---|---|---|---|---|
| ModernBERT-base, fine-tuned | English | all | 0.9790 | 0.9753–0.9822 | 0.9786 |
| ModernBERT-base, fine-tuned | English | 1/2 | 0.9764 | 0.9718–0.9796 | 0.9773 |
| ModernBERT-base, fine-tuned | English | 1/10 | 0.9794 | 0.9771–0.9822 | 0.9770 |
| ModernBERT-base, fine-tuned | English | 1/100 | 0.7092 | 0.6707–0.7491 | 0.5994 |
| ModernBERT-large, fine-tuned | English | all | 0.9853 | 0.9848–0.9856 | 0.9848 |
| ModernBERT-large, fine-tuned | English | 1/2 | 0.9827 | 0.9803–0.9864 | 0.9825 |
| ModernBERT-large, fine-tuned | English | 1/10 | 0.9802 | 0.9795–0.9813 | 0.9770 |
| ModernBERT-large, fine-tuned | English | 1/100 | 0.7461 | 0.6497–0.8164 | 0.6605 |
| Japanese ModernBERT, fine-tuned | Japanese checked | all | 0.9576 | 0.9557–0.9599 | 0.9513 |
| Japanese ModernBERT, fine-tuned | Japanese checked | 1/2 | 0.9458 | 0.9365–0.9536 | 0.9356 |
| Japanese ModernBERT, fine-tuned | Japanese checked | 1/10 | 0.9299 | 0.9270–0.9322 | 0.9138 |
| Japanese ModernBERT, fine-tuned | Japanese checked | 1/100 | 0.6684 | 0.6005–0.7314 | 0.5122 |
| Japanese ModernBERT, fine-tuned | Japanese MT | all | 0.9695 | 0.9677–0.9721 | 0.9679 |
| Japanese ModernBERT, fine-tuned | Japanese MT | 1/2 | 0.9679 | 0.9659–0.9702 | 0.9626 |
| Japanese ModernBERT, fine-tuned | Japanese MT | 1/10 | 0.9545 | 0.9534–0.9564 | 0.9461 |
| Japanese ModernBERT, fine-tuned | Japanese MT | 1/100 | 0.6746 | 0.6100–0.7438 | 0.5230 |
| Japanese ModernBERT, fine-tuned | Japanese revised | all | 0.9611 | 0.9601–0.9625 | 0.9570 |
| Japanese ModernBERT, fine-tuned | Japanese revised | 1/2 | 0.9505 | 0.9443–0.9589 | 0.9412 |
| Japanese ModernBERT, fine-tuned | Japanese revised | 1/10 | 0.9325 | 0.9297–0.9348 | 0.9195 |
| Japanese ModernBERT, fine-tuned | Japanese revised | 1/100 | 0.6724 | 0.6032–0.7384 | 0.5142 |
| Japanese ModernBERT, fine-tuned | External | all | 0.8927 | 0.8656–0.9144 | 0.5711 |
| Japanese ModernBERT, fine-tuned | External | 1/2 | 0.8958 | 0.8684–0.9152 | 0.5613 |
| Japanese ModernBERT, fine-tuned | External | 1/10 | 0.8226 | 0.7428–0.8895 | 0.5243 |
| Japanese ModernBERT, fine-tuned | External | 1/100 | 0.3747 | 0.3577–0.3987 | 0.1261 |

**Table S12. Comparison with prior reports**

| Prior report | Reported | This study | Assessment |
|---|---|---|---|
| Japanese ModernBERT, internal exact match [16] | 0.747 | 0.7444 (mean of three seeds) | Consistent |
| Same, external exact match [16] | 0.5514 | 0.6077 (seed range 0.4856–0.6872) | Reported value within our seed range |
| Same, external mAP [16] | 0.7776 | 0.8156 (seed range 0.8024–0.8404) | Consistent in direction |
| CT-RATE-JPN: CT-BERT-JPN micro-F1 from machine-translated to radiologist-revised text (resident revision followed by board-certified radiologist review; the checked version here) [15] | −0.0175 | Encoder −0.0119 (machine-translated to checked) | Same direction |
| Jev: tuned thresholds improve binary probabilities [9] | Large gains | Jev, 0.5 to development thresholds: +0.1090 external | Consistent |
| Jev: overprediction of "Yes" with independent binary questions [9] | Positive bias | External precision 0.6568–0.8163, recall 0.9648–0.9754 for typed decisions | Consistent |

#### Limitations S1. Further limitations

1. The internal test set is small (150 reports); per-finding F1 and macro-F1 are unstable, with 10 findings having fewer than 30 positives in Japanese.
2. In the reduced-data encoder analysis, each seed determined both the sampled training reports and model training; their separate effects cannot be estimated.
3. Performance was not examined by sex, age or other patient subgroups. CT-RATE comes from one hospital in Türkiye and RR-Findings from 27 Japanese lung cancer cases, so performance and fairness in other populations, institutions and reporting styles are unknown.

#### Methods S2. Data, reference standard, prompts and computing

**Data characteristics.** CT-RATE comprises non-contrast chest CT examinations from one hospital in Türkiye acquired between May 2015 and January 2023 [14]. Table S13 gives the characteristics of each partition from the distributed metadata, and Fig. S1 shows the flow of reports. The 150 internal test reports were randomly sampled from the CT-RATE validation set by the CT-RATE-JPN authors [15]; each comes from a different patient according to the CT-RATE patient identifiers (Table S13). RR-Findings derives from the RR-TNM dataset of the NTCIR-17 MedNLP-SC shared task and provides no demographic data or examination dates [16]. The sizes of the test sets were fixed by the public datasets; no sample size calculation was performed.

**Table S13. Characteristics of the data partitions**

| Characteristic | Training | Development | Development subset | Internal test | External test |
|---|---|---|---|---|---|
| Reports | 20,444 | 2,272 | 450 | 150 | 243 |
| Patients | 16,867 | 1,910 | 426 | 150 | 27 cases |
| Age, median (IQR), years | 48 (36–63) | 47 (36–62) | 47 (36–61) | 47.5 (34–64) | Not available |
| Women, n (%) | 8,420 (41.2%) | 952 (41.9%) | 199 (44.2%) | 63 (42.0%) | Not available |
| Study dates | Jan 2016 – Nov 2022 | Jan 2016 – Nov 2022 | Jan 2016 – Nov 2022 | Nov 2016 – Sep 2022 | Not available |
| Positive labels, % of 18 labels per report | 20.0% | 20.1% | 19.5% | 21.7% | 13.0% |
| Reports with no positive finding | 1,339 | 144 | 32 | 8 | 11 |

Training, development and internal test data are from CT-RATE and CT-RATE-JPN; the external test data are from RR-Findings. Age was missing for 7 training reports and 1 development report, and sex for 3 training reports. Study dates are from the CT-RATE metadata. The internal test labels are the same in English and Japanese.

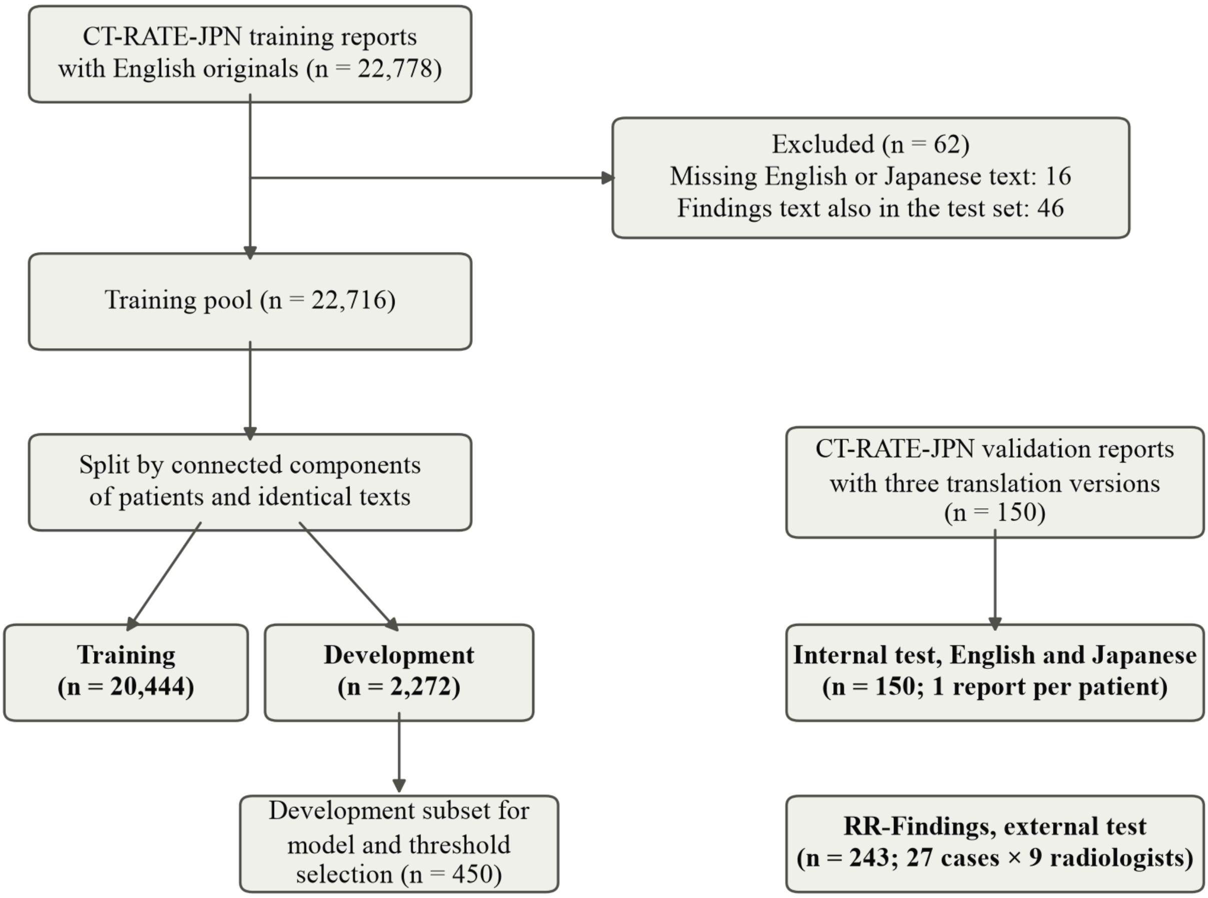


**Fig. S1.** Flow of reports. The development subset was used for encoder selection and threshold fitting; the remaining development reports were not used for selection.

**Reference standard.** CT-RATE labels were produced by RadBERT-RoBERTa-4m fine-tuned on 1,000 reports manually annotated for the 18 findings (800 for training, 200 for validation) and applied to the remaining reports; the labeler's precision, recall and F1 are reported in the CT-RATE publication [14]. CT-RATE-JPN keeps the CT-RATE labels, and none of the 2,700 internal test labels differed between languages. In RR-Findings, a radiology resident (postgraduate year 5) labeled each report using the CT-RATE schema, a board-certified radiologist (postgraduate year 12) reviewed all labels and proposed corrections, and final labels were set by consensus; inter-rater agreement and annotation software were not reported [16]. Three findings have no positive external report (Table S5).

**Input text.** Internal reports were given as "Findings:" followed by the Findings text and "Impressions:" followed by the Impressions text (Japanese headings 所見 and 結論), without truncation or normalization; encoders received the same text. External reports were given as the full report text without change; the instructions, fixed on CT-RATE, still refer to Findings and Impressions. Reports were never truncated: encoder input was limited to 8,192 tokens or the model's maximum length, whichever was smaller, and no report exceeded it.

**Prompt design.** The authors wrote the prompts from the task definition and reviewed them against the published CT-RATE labeling procedure before any test run. No prompt variants were compared on development or test performance. The only rule beyond the task statement, counting suspected or possible findings as present, was a study decision rather than a documented rule of the CT-RATE labeler. The English instructions were:

> Determine which of the specified findings are present according to this CT report. Use only Findings and Impressions. Evaluate each finding independently; an explicitly negated finding is absent. Do not add patient information or use external tools. Treat suspected or possible findings as present, even when they are not definitively confirmed.

The Japanese instructions have the same content; both versions are in `prompts/definition.json` of the code repository.

**Prompts by approach.** Generation: the system message contained the instructions, the output instruction ("A JSON object with exactly the 18 label names as keys and integer 0 (absent) or 1 (present) values.") and "Labels:" followed by the 18 finding names (Japanese: Japanese name with the English name in parentheses); the user message contained the report. Output was constrained by a strict JSON schema with the 18 English finding names as required integer keys taking 0 or 1. Few-shot conditions inserted eight training reports (seed 42; the same cases in both languages, English text or Japanese machine translation) as user and assistant turns with their CT-RATE labels before the target report. Jev, Luna Decisions and Gemini Decisions: the report was the input, and each of the 18 questions consisted of the instructions followed by "Is {finding} present in this report?" (Japanese: 「このレポートに{finding}は認められますか？」). Qwen typed decision: one user message per finding containing the report, the same question and "Answer Yes or No.", with thinking disabled and one output token.

**Inference settings.** Sampling settings are given in Section 2.3. For MedGemma, the model card gives a greedy-decoding example (do_sample=False) but no numerical temperature or top-p recommendation [23]; we therefore used temperature 0 and top-p 1. Maximum output length was 8,192 tokens for Luna, MedGemma and Qwen generation with thinking off, and 16,384 tokens for Qwen with thinking on, except in the first Japanese run of Qwen with thinking on, which used 8,192 tokens and retried one truncated request with 16,384 tokens. Luna requests used the Responses API with storage disabled and no tools. Table S14 lists model versions, access routes, inference dates and knowledge cutoffs.

**Table S14. Models, access and inference dates**

| Model | Conditions | Version | Access | Inference dates (UTC, 2026) | Knowledge cutoff |
|---|---|---|---|---|---|
| Gemini Decision 1 | Gemini Decisions | gemini-decision-1; response revision unavailable | Google AI Studio, v1beta | 22 Nov (local execution records) | Not specified in the cited release [8] |
| gpt-6-luna | Luna generation; Luna Decisions | gpt-6-luna (default snapshot) | OpenAI API | Generation 21 Oct – 4 Nov; Luna Decisions 25 Oct and 29 Oct | 18 May 2026 [20] |
| Jev | Jev | jev-1.14.0 | TypeSafe AI API | 25 Oct – 9 Nov | Not disclosed |
| Qwen3.8-27B | Qwen generation; Qwen typed decision | Qwen/Qwen3.8-27B, Hugging Face revision 1d4bf0f2 | Local, vLLM | 20 Oct – 9 Nov | Not stated in the model card |
| MedGemma | MedGemma generation | google/medgemma-27b-text-it, Hugging Face revision 5b667cf2 | Local, vLLM | 19 Oct – 7 Nov | Not stated in the model card |

Dates cover all test runs used in the tables. Inference dates of Luna generation and local models are from the response timestamps; those of Jev and Luna Decisions, whose responses carry no timestamp, and of Gemini Decisions are from the run logs and output files. All three datasets were publicly available before inference.

**Encoders.** Encoders were initialized from pretrained checkpoints (BioClinical-ModernBERT-base revision c3648aa8, BioClinical-ModernBERT-large revision c684f470, modernbert-ja-130m revision 28c180b1) with a newly initialized 18-output sequence-classification head, and trained with binary cross-entropy loss per finding and AdamW at a constant learning rate without warm-up (other optimizer settings at PyTorch defaults). Each optimizer step accumulated gradients over 8 reports (batch size 1), with bfloat16 mixed precision and no data augmentation. Models from different seeds were evaluated separately and were not ensembled.

**Software and hardware.** Encoder training and local LLM inference used one workstation with two NVIDIA RTX A6000 GPUs (48 GB each) and CUDA 13.0. Training used Python 3.13, PyTorch 2.14.1 and Transformers 5.19.0. Local LLMs were served by vLLM 0.31.0 (PyTorch 2.13.0, Transformers 5.17.0) with tensor parallelism over both GPUs. API conditions were run from a macOS computer with Python 3.13.5, the OpenAI Python SDK 3.26.1 and the TypeSafe SDK 0.7.2; Gemini Decisions used google-genai 3.6.1. Thresholds, metrics, bootstrap intervals and tables were computed with Python and NumPy 2.5.3 using the code in the repository. Encoder training time was not recorded.

#### Probability distributions

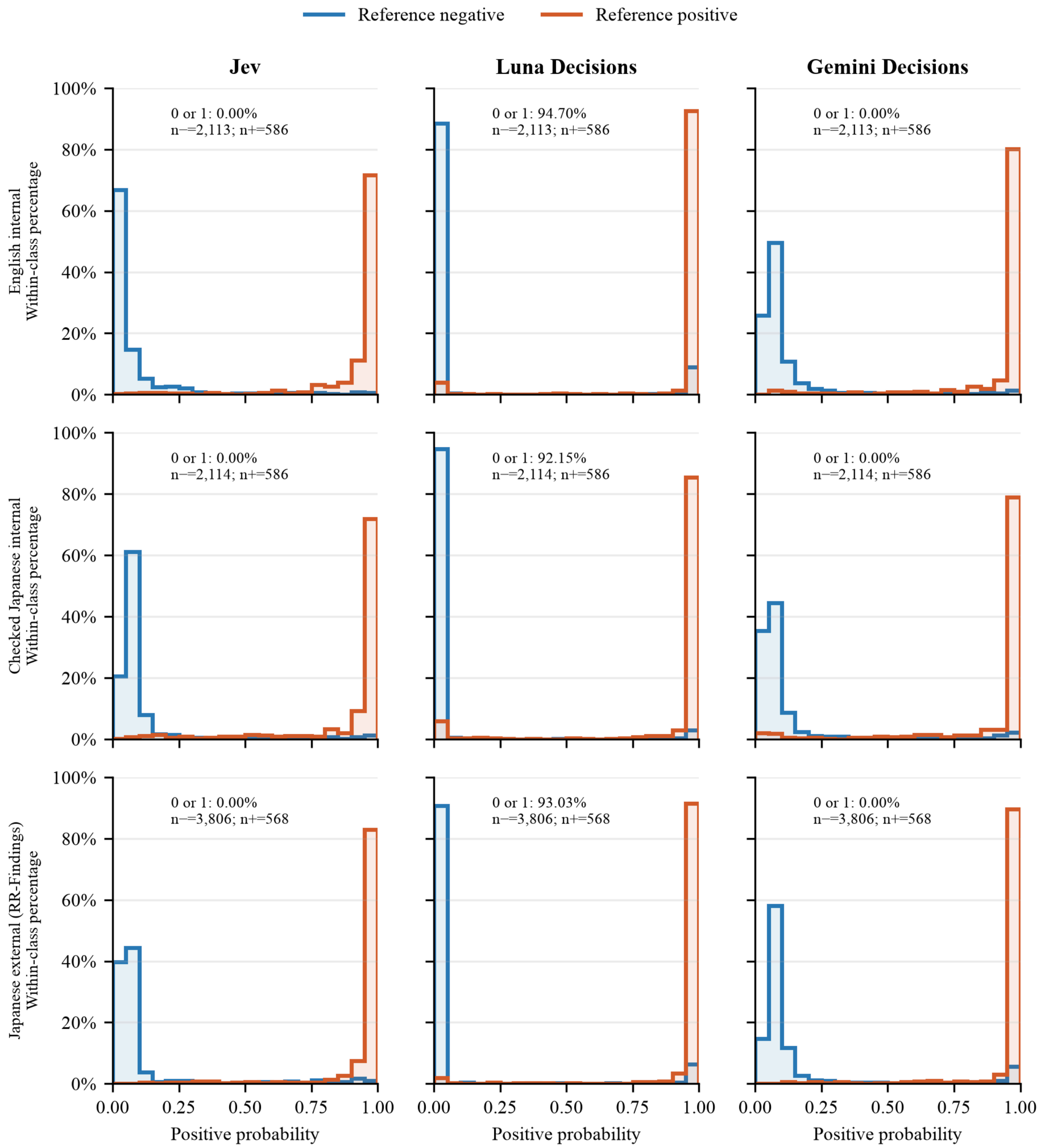


Fig. S2. Returned positive-probability distributions. Rows show English internal, radiologist-checked Japanese internal and naturally written Japanese external reports. Columns show Jev, Luna Decisions and Gemini Decisions. Each panel uses the first adopted run, selected before this analysis. Histograms use common bins of width 0.05; the last bin includes 1. Each reference class is normalized separately to 100%, so the curves do not represent prevalence. All services use the same successfully returned probability cells: 2,699 in English (one Luna Decisions refusal excluded), 2,700 in checked Japanese and 4,374 externally. Missing probabilities were not replaced with zero. The annotations report probabilities exactly equal to 0 or 1 and the negative/positive cell counts. These are descriptive distributions, not calibration curves; the cells are not independent sampling units.
